# Alcohol use disorder accelerates age-related cognitive decline

**DOI:** 10.64898/2026.09.22.26363720

**Authors:** Sarah M. Hartz, Jacquelyn Meyers, Martin H. Plawecki, Kathleen K. Bucholz, Grace Chan, Danielle Dick, Fanghong Dong, Howard J. Edenberg, Christina Garasky, Emma Johnson, Chella Kamarajan, Sivan Kinreich, John R. Kramer, Jodi Kutzner, Alex P. Miller, Zoe Neale, Ashwini Pandey, Gayathri Pandey, Gita Pathak, Jessica E. Salvatore, Marc A. Schuckit, Maggie Clapp Sullivan, Andrey Anokhin, Bernice Porjesz, Laura J. Bierut

**Affiliations:** Washington University in St. Louis; SUNY Downstate Health Sciences University; Indiana University; University of Connecticut; Robert Wood Johnson Medical School; University of Iowa; Icahn School of Medicine at Mount Sinai; UC San Diego

**Author notes:** Disclosures: SMH received speaking fees from Novo Nordisk.

## Abstract

**Background:** Alcohol use disorder (AUD) is an established contributor to neurocognitive impairment and a major modifiable risk factor for dementia. However, the extent to which lifetime AUD interacts with aging to accelerate subtle cognitive trajectories prior to overt clinical dementia remains unclear.

**Objective:** To investigate whether a lifetime history of DSM-5 AUD and correlated indices of alcohol consumption are associated with accelerated age-related cognitive decline in middle-aged and older adults.

**Methods:** Participants (N = 1,421) were drawn from the Collaborative Study on the Genetics of Alcoholism (COGA) prospective longitudinal study. Lifetime AUD status (defined as 2 or more DSM-5 criteria endorsed at peak drinking) and drinking patterns were derived from the Semi-Structured Assessment for the Genetics of Alcoholism (SSAGA). Cognitive performance across multiple domains was evaluated using the NIH Toolbox Cognition Battery to yield Total, Fluid, and Crystallized Composite Scores. Linear mixed-effects models examined main effects of AUD and age, as well as an AUD-by-age interaction, controlling for testing site.

**Results:** At the time of cognitive assessment, the overall sample had a mean age of 52.5 years (SD = 14.5) with a mean longitudinal observation period of 25.0 years (SD = 4.7). The sample was 60% female, 40% male, 7% Hispanic, 23% non-Hispanic Black, 68% non-Hispanic White, 2% other race/ethnicity. 64% met lifetime criteria for AUD (n = 908). While baseline mean cognitive composite scores were similar between those with and without AUD, lifetime AUD was significantly associated with steeper age-related decline for both the Total Composite score (interaction estimate = −0.092, 95% CI [−0.173, −0.010], p = 0.03) and Fluid Composite score (interaction estimate = −0.13, 95% CI [−0.220, −0.040], p = 0.005). No significant AUD-by-age interaction was observed for Crystallized Composite scores (interaction estimate = −0.026, 95% CI [−0.092, 0.041], p = 0.449). Based on model projections, the AUD-associated cognitive deficit on Total Composite scores was equivalent to 7.1 additional years of cognitive aging at age 60 (95% CI [−0.5, 14.7]) and 13.8 additional years at age 75 (95% CI [0.2, 27.3]). Measures of current (past-year) drinking status and current drinks per week showed no significant interaction with age.

**Conclusions:** A lifetime history of AUD acts as a catalyst for cognitive aging, significantly steepening the trajectory of age-related decline in total and fluid cognitive functioning well before the onset of clinical dementia. Cross-sectional measures of current drinking failed to capture these effects, demonstrating that evaluating cumulative lifetime drinking history is important for assessing alcohol-related cognitive decline.

## Introduction

Alcohol use disorder (AUD) significantly contributes to cognitive decline and is increasingly recognized as a major modifiable risk factor for neurodegenerative conditions, including Alzheimer’s disease (AD).^1^ Excessive alcohol consumption exerts severe neurotoxic effects on the aging brain through mechanisms such as thiamine deficiency, neuroinflammation, oxidative stress, and structural brain atrophy.^2,3^ Because rates of alcohol consumption among middle-aged and older adults have risen substantially over recent decades, the population-level burden of alcohol-related cognitive decline and secondary neurodegenerative disorders is projected to increase.^4^

The clinical association between heavy, chronic alcohol consumption and severe, irreversible cognitive impairment has been established for decades.^5^ The most well-documented manifestation of severe alcohol-induced neurotoxicity is Wernicke-Korsakoff syndrome (WKS)—a neurological disorder resulting from alcohol-induced thiamine (vitamin B1) deficiency.^6^ While Wernicke’s encephalopathy presents acutely with confusion, ataxia, and ophthalmoplegia, its progression into Korsakoff’s psychosis results in profound persistent anterograde amnesia, executive dysfunction, and confabulation due to damage within the diencephalon and limbic structures.^7,8^ Beyond classic nutritional deficiencies, chronic heavy drinking directly induces frontostriatal and cerebellar atrophy, leading to widespread cognitive deficits even in the absence of a formal WKS diagnosis.^9,10^

While major neurocognitive end-points—such as clinical dementia diagnoses and alcohol-related brain damage—are crucial clinical markers, focusing exclusively on these extreme endpoints overlooks earlier, subtle stages of cognitive impairment. Epidemiological studies consistently show that AUD and heavy drinking (typically defined as >14 drinks per week) double to triple the risk of developing dementia in later life.^3,11,12^ Furthermore, emerging prospective studies suggest that AUD accelerates subtle cognitive trajectories years prior to clinical dementia onset.^13^ These cognitive effects associated with lower levels of alcohol consumption may effect a larger segment of the population. However, a key question remains regarding the magnitude of AUD-associated cognitive decline in older populations interacts with the aging process.^14^

This study investigates whether a lifetime history of DSM5 AUD and correlated measures of alcohol consumption such as maximum alcoholic drinks consumed are associated with accelerated cognitive decline in middle-aged and older adults prior to the onset of overt clinical dementia.

## Methods

### Participants

The Collaborative Study on the Genetics of Alcoholism (COGA) is a family-based longitudinal study that was initiated in 1989 to understand the development of alcohol use disorder and its consequences.^15^ Established in 1990 as a high-risk family study, COGA initially recruited probands in treatment for Alcohol Use Disorder (AUD) alongside their first-degree relatives and community comparison families across six sites. The Collaborative Study on the Genetics of Alcoholism (COGA) target lifespan sample comprises 6,322 well-characterized participants aged 40 and older selected from the broader prospective, family-based COGA pool (N = 17,885 across 2,246 families). The COGA lifespan cohort used in this study consists of N=1451 COGA participants who were at least 40 years of age at the most recent COGA assessment between 2020 and 2024, and had at least one prior baseline assessment. All participants provided informed consent prior to participation.

### Assessments

The Semi-Structured Assessment for the Genetics of Alcoholism (SSAGA) is the longitudinal assessment battery and was used to extract baseline and longitudinal information regarding demographics and longitudinal alcohol use.^16,17^ Cognitive functioning was measured using the NIH-Toolbox Cognitive Batteries.^18^ This battery consists of tests of multiple cognitive constructs that measure attention, executive function, language, memory, learning, and processing speed: Dimensional Change Card Sort, Flanker Inhibitory Control and Attention, List Sorting Working Memory, Oral Reading Recognition, Pattern Comparison, Picture Sequence Memory, and Picture Vocabulary Recognition.

### Alcohol related variables

Lifetime history of AUD was used to reflect cumulative alcohol-related burden and alignment with prior work on long-term neuropathological effects, and was defined based on having 2 or more DSM-5-TR^19^ AUD symptoms at the time of participants’ heaviest lifetime drinking, assessed during the SSAGA interviews.^16,17^ Current alcohol consumption was calculated using self-reported drinking quantities also collected as part of the SSAGA. Participants reported the average number of standard drinks consumed per week, based on typical drinking patterns during the most recent period of alcohol use. These values were used to derive a continuous variable representing current drinks per week.

### Measures of cognition

Using a demographically-corrected standardized scoring algorithm, the NIH-toolbox battery was compiled into a total composite score, a fluid composite score, and a crystallized composite score.^20^

### Statistical Analyses

All statistical analyses were performed using SAS 9.4 or R version 4.6.0.^21,22^ Statistical significance was defined at an alpha level of 0.05. We used linear regression to evaluate the association between the cognitive composite scores, age and the alcohol use variables, focusing on the interaction between the age and alcohol use variable. A random effect for the COGA testing site was included.

## Results

The primary analytical sample comprised 1,421 participants with a mean longitudinal observation period of 25.0 years (SD = 4.7). At the time of cognitive assessment, the mean age across the sample was 52.5 years (SD = 14.5). **Table 1** summarizes the demographic composition, alcohol consumption patterns, diagnostic profiles, and baseline cognitive performance of the cohort. The overall cohort was 60% female and 40% male. Self-reported racial and ethnic composition was 68% non-Hispanic White, 23% non-Hispanic Black, 7% Hispanic, and 2% other race/ethnicity. A lifetime history of DSM-5 AUD, defined as endorsing two or more criteria during peak drinking, was present in 64% of the sample. Among participants with lifetime AUD, severe presentation was common: 22% (n = 204) endorsed 9 or more AUD symptoms at peak severity, 18% (n = 164) endorsed 6 to 8 symptoms, and 21% (n = 189) endorsed 4 to 5 symptoms. Peak 24-hour alcohol consumption was markedly elevated among participants with AUD (median maximum drinks = 20) compared to those without AUD (median maximum drinks = 7). At the time of assessment, 75% (n = 1,072) of the total sample were current past-year drinkers, while 23% (n = 329) were former drinkers, and 1.4% (n = 20) were never drinkers. Current drinks per week averaged 6.7 (SD = 13.7) across the full cohort, with higher average weekly consumption observed among individuals with lifetime AUD (9.0 drinks per week, SD = 16.4) relative to those without AUD (2.5 drinks per week, SD = 3.0). Unadjusted mean NIH Toolbox standardized scores were similar across participants with and without AUD for the Total Composite (100.9 vs. 102.1), Fluid Composite (97.3 vs. 99.0), and Crystallized Composite (105.3 vs. 105.6) scores.

**Table 1.** Participant characteristics.

|  | **Overall** | **No AUD** | **AUD** |
| --- | --- | --- | --- |
|  | N=1421 | N = 513 | N = 908 |
| **Age at cognitive assessment** | 52.5 (14.5) | 51.7 (15.8) | 53.0 (13.7) |
| **Years observed** | 25.0 (4.7) | 24.6 (4.9) | 25.3 (4.5) |
| **Sex** |  |  |  |
| Male | 570 (40%) | 142 (28%) | 428 (47%) |
| Female | 851 (60%) | 371 (72%) | 480 (53%) |
| **Race/ethnicity** |  |  |  |
| Black, non-Hispanic | 326 (23%) | 132 (26%) | 194 (21%) |
| Hispanic | 99 (7.0%) | 26 (5.1%) | 73 (8.0%) |
| White, non-Hispanic | 970 (68%) | 344 (67%) | 626 (69%) |
| Other race/ethnicity | 26 (1.8%) | 11 (2.1%) | 15 (1.7%) |
| **Alcohol use** |  |  |  |
| Current drinker (past year) | 1,072 (75%) | 371 (72%) | 701 (77%) |
| Former drinker | 329 (23%) | 122 (24%) | 207 (23%) |
| Never drinker | 20 (1.4%) | 20 (3.9%) | 0 (0%) |
| **Current drinks per week** | 6.7 (13.7) | 2.5 (3.0) | 9.0 (16.4) |
| **Number of AUD criteria endorsed** |  |  |  |
| 0 | 307 (22%) | 307 (60%) | 0 (0%) |
| 1 | 206 (14%) | 206 (40%) | 0 (0%) |
| 2 | 176 (12%) | 0 (0%) | 176 (19%) |
| 3 | 175 (12%) | 0 (0%) | 175 (19%) |
| 4-5 | 189 (13%) | 0 (0%) | 189 (21%) |
| 6-8 | 164 (12%) | 0 (0%) | 164 (18%) |
| 9+ | 204 (14%) | 0 (0%) | 204 (22%) |
| **Cognitive test measures** |  |  |  |
| Total Composite | 101.3 (12.2) | 102.1 (12.5) | 100.9 (12.0) |
| Fluid Composite | 97.9 (14.7) | 99.0 (14.7) | 97.3 (14.7) |
| Crystallized Composite | 105.4 (9.5) | 105.6 (10.0) | 105.3 (9.2) |

Mean (SD); n (column %)

We ran linear mixed-effects models with the cognitive outcomes and individual alcohol-related variables, including an interaction with age, incorporating random study-site effects. The full model parameter estimates are given in **Supplementary Table 1**. Age at testing was strongly and significantly associated with cognitive performance across all three NIH Toolbox composite measures. Lifetime history of AUD significantly interacted with age to accelerate the trajectory of age-related cognitive decline, as detailed in **Table 2**. For the Total Composite score, lifetime AUD was associated with a significantly steeper slope of age-related decline (interaction beta = −0.092, 95% CI [−0.173, −0.010], p = 0.03). The accelerating effect of lifetime AUD was even more pronounced for the Fluid Composite score (interaction beta = −0.130, 95% CI [−0.220, −0.040], p = 0.005). In contrast, lifetime AUD did not significantly interact with age for Crystalized Composite score (interaction beta = −0.026, 95% CI [−0.092, 0.041], p = 0.449). **Figure 1** illustrates these differential trajectories across age: while predicted cognitive scores for individuals with and without AUD are comparable in midlife around age 50, the trajectories diverge progressively across older ages. The divergence is steepest for the Fluid Composite domain, intermediate for the Total Composite domain, and virtually absent for the Crystallized Composite domain.

**Table 2.** PEstimated association of alcohol use on age-related cognition. Estimated interaction regression parameter from linear regression models for cognitive outcomes including age at testing, alcohol parameter, and the interaction between the two. Models also include a random effect for the COGA study site. Table includes model parameters corresponding to the interaction effect. All model parameter estimates are given in Supplementary Tables.

| Alcohol variable interaction with age |  | Total Composite | Fluid Composite | Crystallized Composite |
| --- | --- | --- | --- | --- |
| Lifetime alcohol use disorder | Estimate | -0.092 | -0.13 | -0.026 |
|  | 95% CI | [-0.173, -0.010] | [-0.220, -0.040] | [-0.092, 0.041] |
|  | p-value | 0.03 | 0.005 | 0.449 |
| Number of alcohol use disorder symptoms | Estimate | -0.014 | -0.017 | -0.006 |
|  | 95% CI | [-0.027, -0.000] | [-0.032, -0.002] | [-0.017, 0.005] |
|  | p-value | 0.047 | 0.02 | 0.28 |
| Lifetime max drinks in 24h | Estimate | -0.008 | -0.009 | -0.005 |
|  | 95% CI | [-0.01, -0.002] | [-0.016, -0.002] | [-0.010, 0.000] |
|  | p-value | 0.01 | 0.01 | 0.055 |
| Current vs. former drinker (among ever drinkers) | Estimate | -0.082 | -0.086 | -0.053 |
|  | 95% CI | [-0.182, 0.017] | [-0.196, 0.024] | [-0.134, 0.028] |
|  | p-value | 0.105 | 0.127 | 0.2 |
| Drinks per week | Estimate | -0.001 | -0.001 | -0.001 |
|  | 95% CI | [-0.005, 0.002] | [-0.005, 0.003] | [-0.004, 0.002] |
|  | p-value | 0.48 | 0.544 | 0.59 |

**Figure 1.**
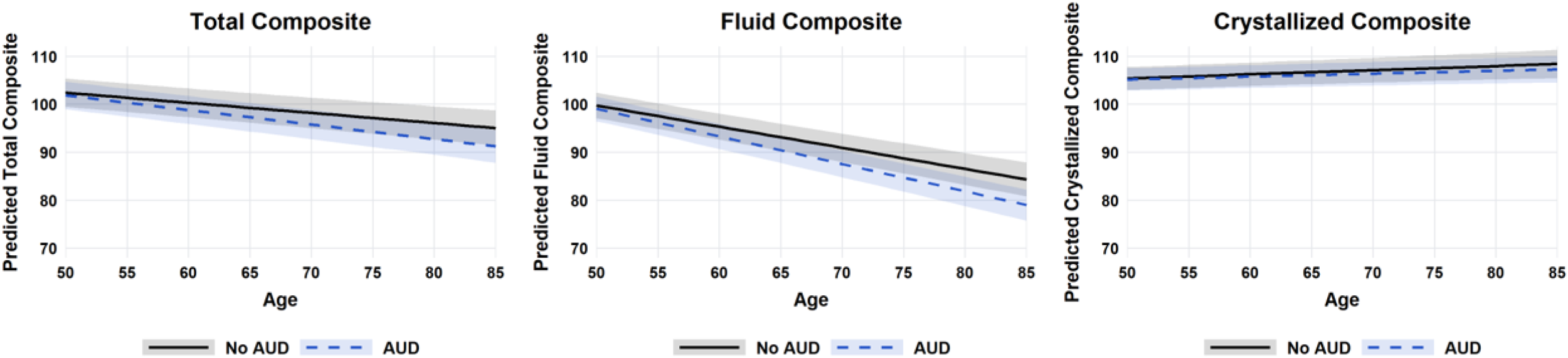
Plots of estimated cognitive score by age, based on regression model estimates from Table 2.

Similar age-related associations with cognitive decline were seen for symptom count and for the effect of maximum drinks in a 24-hour period (max drinks). Specifically, endorsement of a higher number of lifetime AUD symptoms was significantly associated with steeper age-related decline for both the Total Composite (interaction beta = −0.014, 95% CI [−0.027, 0.000], p = 0.047; **Table 2**) and Fluid Composite (interaction beta = −0.017, 95% CI [−0.032, −0.002], p = 0.02) domains. The maximum number of drinks consumed within a 24-hour period similarly predicted accelerated age-related decline in Total Composite scores (interaction beta = −0.008, 95% CI [−0.014, −0.002], p = 0.01) and Fluid Composite scores (interaction beta = −0.009, 95% CI [−0.016, −0.002], p = 0.01). In contrast, cross-sectional measures of past-year drinking failed to demonstrate significant interactions with age. Neither current versus former drinker status (Total Composite interaction beta = −0.082, p = 0.105) nor continuous current drinks per week (Total Composite interaction beta = −0.001, p = 0.48) significantly moderated age-related cognitive trajectories.

To contextualize the clinical magnitude of the lifetime AUD effect on overall cognitive trajectory, model-derived estimates of AUD-attributable cognitive deficit were converted into equivalent years of cognitive aging as a function of age in **Table 3**. Because the AUD-by-age interaction compounds over time, the cognitive deficit attributable to lifetime AUD increases substantially across advancing age brackets. At age 55, lifetime AUD is equivalent to 4.9 years of additional cognitive aging (95% CI [−1.4, 11.2]). At age 60, lifetime AUD is equivalent to 7.1 years of additional cognitive aging (95% CI [−0.5, 14.7]). At age 65, this effect becomes statistically significant and corresponds to 9.3 years of additional cognitive aging (95% CI [0.0, 18.7]). Finally, at age 75, the cognitive impact of lifetime AUD on Total Composite function reaches an equivalent of 13.8 years of additional cognitive aging (95% CI [0.2, 27.3]).

**Table 3.** Estimated equivalent cognitive years of aging from AUD, as a function of current age. Estimates are based on the total composite score. For example, those with AUD at age 65 are estimated to have the same cognition as someone without AUD who is 9 years older (age 74).

| Baseline Age | Estimated cognitive aging equivalent to AUD [95% CI] |
| --- | --- |
| 45 | 0.5 [-5.9, 6.9] |
| 50 | 2.7 [-3.2, 8.6] |
| 55 | 4.9 [-1.4, 11.2] |
| 60 | 7.1 [-0.5, 14.7] |
| 65 | 9.3 [0.0, 18.7] |
| 70 | 11.5 [0.2, 22.9] |
| 75 | 13.8 [0.2, 27.3] |

## Discussion

Our findings demonstrate that AUD significantly accelerates cognitive decline associated with aging. Heavy alcohol consumption appears to act as a catalyst, steepening the rate of cognitive deterioration over time in older cohorts. For example, at age 60, the impact of AUD on cognitive functioning is equivalent to 7 years of aging and at age 75, the impact of AUD on cognitive functioning is equivalent to 14 years of aging.

These results align with and extend a growing body of epidemiological and clinical literature linking heavy alcohol consumption to adverse neurocognitive outcomes.^1,13^ While landmark population studies have established AUD as a major modifiable risk factor for end-stage dementia,^12^ our study highlights that the burden of AUD is evident much earlier along the continuum of cognitive aging, prior to the onset of dementia. Classical frameworks have often focused on severe, acute manifestations of alcohol-related brain damage— such as Wernicke-Korsakoff syndrome.^7,8^ However, our findings confirm that a diagnosis of AUD exerts a more pervasive, insidious effect across middle-aged and older individuals by accelerating age-related cognitive decline prior to the clinical onset of overt neurodegenerative syndromes.

Several distinct, intersecting neurobiological mechanisms may explain how AUD accelerates cognitive aging. First, chronic ethanol exposure directly induces neurotoxicity, leading to structural brain changes including frontal lobe atrophy, loss of white matter integrity, and reduced synaptic density.^3,9,10^ Second, excessive alcohol intake exacerbates systemic and central neuroinflammation, oxidative stress, and cerebrovascular dysfunction, all of which compromise neural resilience in the aging brain.^23,24^ Third, AUD may interact synergistically with primary neurodegenerative pathologies, such as Alzheimer’s disease amyloid and tau burden, lowering the threshold at which pathological protein accumulation manifests as clinically measurable cognitive impairment.^25^

Our study highlights the difficulty of studying the effect of heavy alcohol consumption and AUD on cognitive functioning. Heavy alcohol consumption can be episodic and so having a cross-sectional one-time measure of current (past year) alcohol use may not capture the full picture of a person’s history of alcohol use. For instance, if we used current alcohol consumption as a predictor of cognitive decline, we see no association with cognitive measures. Being able to capture a cumulative lifetime “dosage” of alcohol consumption is critical to better understanding the effects of alcohol on the aging brain. Additionally, age at when alcohol is consumed may be important. A large amount of alcohol consumed as a teenager may have a different long term effect than a similar amount consumed in middle age. Older adults are likely uniquely vulnerable to the neurotoxic effects of alcohol due to age-related changes in ethanol pharmacokinetics, increased blood-brain barrier permeability, and decreased physiological reserve and neuroplasticity.

### Clinical and Public Health Implications

As our population ages and alcohol consumption among middle-aged and older adults continues to rise globally,^4^ these findings carry urgent public health implications. Identifying AUD and heavy drinking as a potent accelerator of cognitive decline underscores the necessity of early screening and targeted intervention in primary care and geriatric settings. Given that alcohol use is a modifiable behavior, addressing AUD in midlife and earlier represents a crucial window for mitigating the AUD associated accelerated cognitive decline and the many resulting consequences that come from aging.

## Data Availability

All data produced in the present study are available upon reasonable request to the authors

## Supplementary Tables

Regression parameters for statistical models

**Supplementary Table 1.** NIH Toolbox Composite score ~ age at testing + lifetime AUD +lifetime AUD * age at testing + random effect for COGA site.

| Outcome Variable | Intercept |  |  | Age |  |  | AUD |  |  | Age x AUD |  |  |
| --- | --- | --- | --- | --- | --- | --- | --- | --- | --- | --- | --- | --- |
| | $\beta$ | 95% CI | p | $\beta$ | 95% CI | p | $\beta$ | 95% CI | p | $\beta$ | 95% CI | p |
| Total Composite | 113.0 | [108.7, 117.4] | 0 | -0.2 | [-0.3, -0.2] | 0 | 4.0 | [-0.5, 8.4] | 0.08 | -0.09 | [-0.17, -0.01] | 0.03 |
| Fluid Composite | 122.0 | [117.6, 126.4] | 0 | -0.4 | [-0.5, -0.4] | 0 | 5.7 | [0.8, 10.6] | 0.02 | -0.13 | [-0.22, -0.04] | 0.005 |
| Crystallized Composite | 101.1 | [97.5, 104.6] | 0 | 0.1 | [0.04, 0.1] | 0.001 | 1.1 | [-2.6, 4.7] | 0.56 | -0.03 | [-0.09, 0.04] | 0.45 |

**Supplementary Table 2.** NIH Toolbox Composite score ~ age at testing + AUD symptom count + AUD symptom count * age at testing + random effect for COGA site.

| Outcome Variable | Intercept |  |  | Age |  |  | AUD symptom count |  |  | Age x AUD symptom count |  |  |
| --- | --- | --- | --- | --- | --- | --- | --- | --- | --- | --- | --- | --- |
| | $\beta$ | 95% CI | p | $\beta$ | 95% CI | p | $\beta$ | 95% CI | p | $\beta$ | 95% CI | p |
| Total Composite | 114.2 | [110.2, 118.3] | 0.0 | -0.2 | [-0.3, -0.2] | 0.00 | 0.28 | [-0.5, 1.0] | 0.47 | -0.014 | [-0.03, 0.00] | 0.05 |
| Fluid Composite | 123.7 | [119.7, 127.8] | 0.0 | -0.5 | [-0.5, -0.4] | 0.00 | 0.42 | [-0.4, 1.3] | 0.33 | -0.017 | [-0.03, -0.002] | 0.02 |
| Crystallized Composite | 101.3 | [98.0, 104.6] | 0.0 | 0.1 | [0.05, 0.14] | 0.00 | 0.06 | [-0.6, 0.7] | 0.84 | -0.006 | [-0.02, 0.01] | 0.28 |

**Supplementary Table 3.** NIH Toolbox Composite score ~ age at testing + max drinks in 24h +max drinks in 24h* age at testing + random effect for COGA site.

| Outcome Variable | Intercept |  |  | Age |  |  | Max drinks |  |  | Age x max drinks |  |  |
| --- | --- | --- | --- | --- | --- | --- | --- | --- | --- | --- | --- | --- |
| | $\beta$ | 95% CI | p | $\beta$ | 95% CI | p | $\beta$ | 95% CI | p | $\beta$ | 95% CI | p |
| Total Composite | 111.5 | [105.7, 117.2] | 0 | -0.2 | [-0.3, -0.1] | 0 | 0.3 | [-0.03, 0.7] | 0.075 | -0.008 | [-0.014, -0.002] | 0.01 |
| Fluid Composite | 120.7 | [114.6, 126.8] | 0 | -0.4 | [-0.5, -0.3] | 0 | 0.4 | [-0.02, 0.7] | 0.064 | -0.009 | [-0.016, -0.002] | 0.01 |
| Crystallized Composite | 99.6 | [94.9, 104.3] | 0 | 0.1 | [0.06, 0.2] | 0 | 0.13 | [-0.1, 0.5] | 0.21 | -0.005 | [-0.010, 0.000] | 0.055 |

**Supplementary Table 4.** NIH Toolbox Composite score ~ age at testing + current/former drinker +current/former drinker* age at testing + random effect for COGA site.

|  | Intercept | Age | Current drinker | Age x current drinker |
| --- | --- | --- | --- | --- |

| Outcome Variable | $\beta$ | 95% CI | p | $\beta$ | 95% CI | p | $\beta$ | 95% CI | p | $\beta$ | 95% CI | p |
| --- | --- | --- | --- | --- | --- | --- | --- | --- | --- | --- | --- | --- |
| Total Composite | 115.0 | [111.3, 118.7] | 0.0 | -0.3 | [-0.3, -0.2] | 0.00 | 0.05 | [-0.1, 0.2] | 0.64 | -0.001 | [-0.005, 0.002] | 0.48 |
| Fluid Composite | 124.9 | [121.3, 128.5] | 0.0 | -0.5 | [-0.6, -0.5] | 0.00 | 0.05 | [-0.1, 0.3] | 0.63 | -0.001 | [-0.005, 0.003] | 0.54 |
| Crystallized Composite | 101.6 | [98.6, 104.6] | 0.0 | 0.1 | [0.04, 0.11] | 0.00 | 0.02 | [-0.1, 0.2] | 0.85 | -0.001 | [-0.004, 0.002] | 0.59 |

**Supplementary Table 5.** NIH Toolbox Composite score ~ age at testing + current DPW+ current DPW * age at testing + random effect for COGA site.

| Outcome Variable | Intercept |  |  | Age |  |  | Current DPW |  |  | Age x current DPW |  |  |
| --- | --- | --- | --- | --- | --- | --- | --- | --- | --- | --- | --- | --- |
| | $\beta$ | 95% CI | p | $\beta$ | 95% CI | p | $\beta$ | 95% CI | p | $\beta$ | 95% CI | p |
| Total Composite | 106.3 | [100.6, 112.1] | 0.0 | -0.2 | [-0.3, -0.08] | 0.00 | 9.4 | [3.7, 15.2] | 0.00 | -0.08 | [-0.18, 0.02] | 0.11 |
| Fluid Composite | 115.8 | [109.7, 121.9] | 0.0 | -0.4 | [-0.5, -0.3] | 0.00 | 10.0 | [3.6, 16.3] | 0.00 | -0.09 | [-0.196, 0.024] | 0.13 |
| Crystallized Composite | 96.0 | [91.3, 100.7] | 0.0 | 0.13 | [0.06, 0.20] | 0.00 | 5.9 | [1.3, 10.6] | 0.01 | -0.05 | [-0.13, 0.03] | 0.20 |

## Notes

Funding: This study was funded by NIH grants R01 AG065234, R01 AG084723, and P01 AG026276 from the National Institute on Aging (NIA), and R01 AA029308 and U10 AA008401 from the National Institute on Alcohol Abuse and Alcoholism (NIAAA). Additional support was provided by the Washington University Institute of Clinical and Translational Sciences through grant UL1TR002345 from the National Center for Advancing Translational Sciences (NCATS) of the NIH.

### Competing Interest Statement

This study was funded by NIH grants R01 AG065234, R01 AG084723, and P01 AG026276 from the National Institute on Aging (NIA), and R01 AA029308 and U10 AA008401 from the National Institute on Alcohol Abuse and Alcoholism (NIAAA). Additional support was provided by the Washington University Institute of Clinical and Translational Sciences through grant UL1TR002345 from the National Center for Advancing Translational Sciences (NCATS) of the NIH. SMH received speaking fees from Novo Nordisk.

### Author Declarations

The IRB of Washington University gave ethical approval for this work.

